# Patterns in Repeat Reinfections: Pre and Post Omicron Emergence

**DOI:** 10.1101/2023.06.29.23292041

**Authors:** Hiam Chemaitelly, Houssein H. Ayoub, Patrick Tang, Peter Coyle, Hadi M. Yassine, Asmaa A. Al Thani, Mohammad R. Hasan, Zaina Al-Kanaani, Einas Al-Kuwari, Andrew Jeremijenko, Anvar H. Kaleeckal, Ali N. Latif, Riyazuddin M. Shaik, Hanan F. Abdul-Rahim, Gheyath K. Nasrallah, Mohamed G. Al-Kuwari, Adeel A. Butt, Hamad E. Al-Romaihi, Mohamed H. Al-Thani, Abdullatif Al-Khal, Roberto Bertollini, Laith J. Abu-Raddad

## Abstract

Our understanding of SARS-CoV-2 reinfection patterns remains limited. We conducted a longitudinal study using Qatar’s national SARS-CoV-2 data from February 28, 2020 to June 11, 2023 to investigate incidence of reinfections both prior to and after omicron emergence. The latter analysis excluded individuals with pre-omicron infections. Before omicron introduction, the proportion of incident infections classified as reinfections gradually increased but remained minimal, reaching 1.8% just before omicron emerged. During the first omicron wave, this proportion reached 9.0%, a 5-fold increase. After the conclusion of the first omicron wave, the proportion of incident infections identified as reinfections rapidly increased, reaching 43.3% towards the end of the study. In the pre-omicron era, a total of 3,131 reinfections were documented, of which 99.6% were first reinfections and 0.4% were second reinfections. Meanwhile, a total of 20,962 reinfections were documented after an omicron primary infection of which 99.0% were first reinfections, 1.0% were second reinfections, and 0.01% were third reinfections. Reinfections were rare before omicron’s emergence but became widespread during the omicron era, including among individuals previously infected with omicron. Our findings may indicate accelerated viral evolution in the omicron era aimed at evading population immunity, but with minimal impact on COVID-19 severity, or potentially suggest immune imprinting effects that require further investigation.

## Main text

Understanding of the pattern of repeat SARS-CoV-2 reinfections remains limited. We conducted a longitudinal study on the entire population of Qatar, investigating the incidence of reinfections both prior to and after omicron emergence.

We derived SARS-CoV-2 laboratory testing, clinical infection, and demographic data from Qatar’s national database, which recorded every SARS-CoV-2 test since the pandemic onset (Sections S1-S2 of Supplementary Appendix). Before November 1, 2022, ∼5% of the population per week underwent testing for SARS-CoV-2 infection, primarily for non-clinical purposes. However, starting from November 1, 2022, the testing rate was reduced to ∼1% per week. This extensive testing approach increases the likelihood of identifying reinfections, regardless of symptomatic presentation. Consequently, this setting presents an exceptional opportunity to analyze patterns of reinfections and repeated reinfections.

Reinfection was defined as an infection documented ≥90 days after a prior infection.^1^ We analyzed data from February 28, 2020 (date of first infection), to June 11, 2023, examining the daily proportions of primary infections, first reinfections, second reinfections, and third reinfections among all incident infections. The analysis was also conducted for two distinct time periods: before omicron introduction on December 1, 2021, and after its introduction, starting from December 19, 2021, coinciding with the first omicron wave.^1^ The latter analysis exclusively examined omicron infections, after excluding individuals with pre-omicron infections. Consequently, all infections analyzed were omicron infections, and all reinfections were subsequent to a primary omicron infection.

Figure 1A illustrates proportion of primary infections and reinfections observed throughout the study, with different variants dominating at different times (Section S3). Prior to omicron introduction, proportion of incident infections classified as reinfections gradually increased but remained minimal, reaching 1.8% just before omicron emerged. However, during the first omicron wave, this proportion experienced a 5-fold surge, soaring to 9.0%. Subsequently, after the conclusion of the first omicron wave, proportion of incident infections identified as reinfections rapidly increased, reaching 43.3% towards the end of the study.

**Figure 1.**
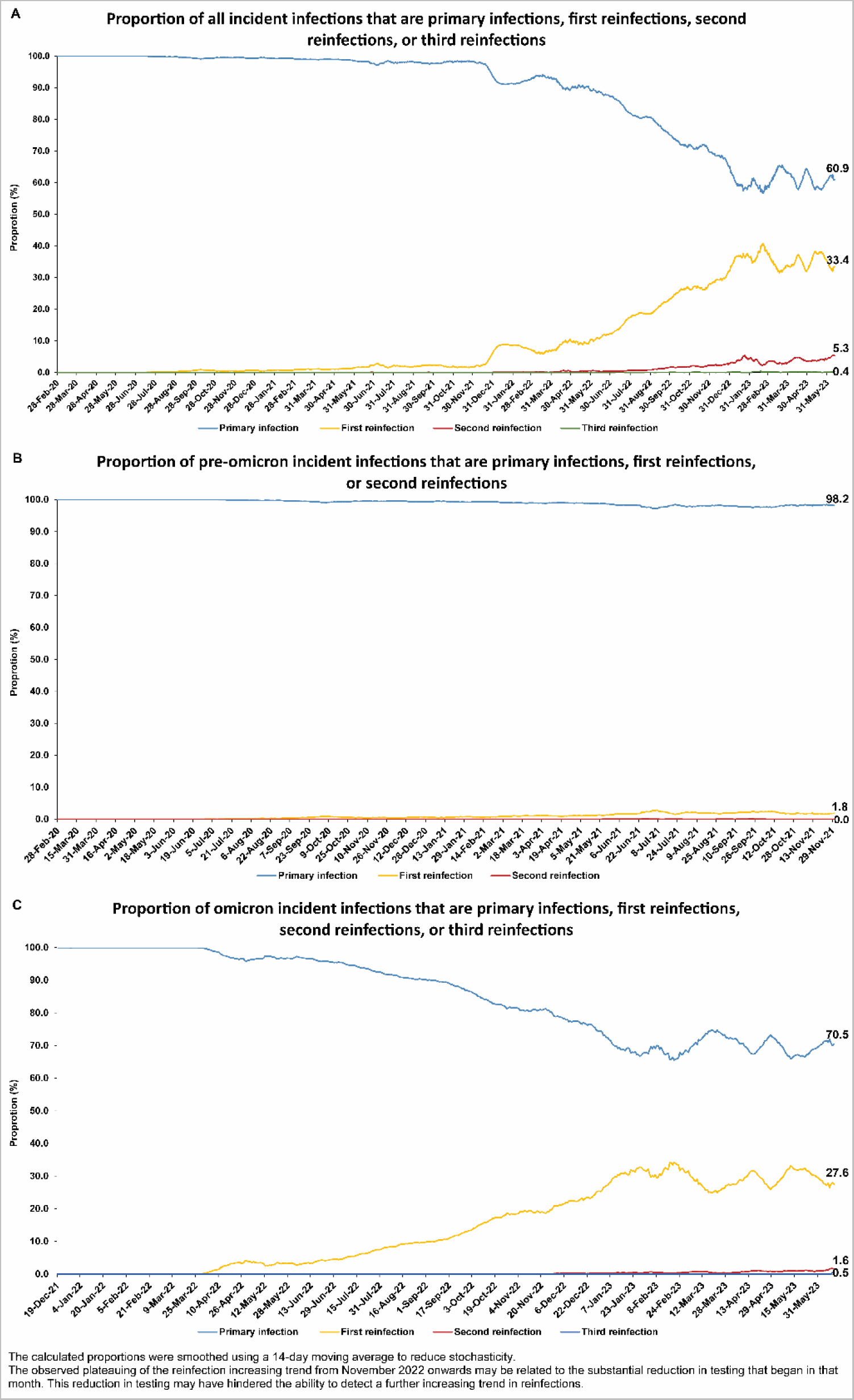
Proportions of A) all incident infections, B) pre-omicron incident infections, and C) omicron incident infections classified as primary infections, first reinfections, second reinfections, or third reinfections. The analysis in C excluded individuals with pre-omicron infections, meaning that all reinfections are subsequent to a primary omicron infection.

In the pre-omicron era (Figure 1B), a total of 3,131 reinfections were documented. Among them, 3,117 (99.6%) were first reinfections, while 14 (0.4%) were second reinfections (Table S1). No cases of third or higher order reinfections were documented.

In the omicron era (Figure 1C), a total of 20,962 reinfections were documented after an omicron primary infection (Table S2). Among these reinfections, 20,744 (99.0%) were first reinfections, 215 (1.0%) were second reinfections, and 3 (0.01%) were third reinfections. No cases of fourth or higher order reinfections were documented.

The patterns of reinfection and repeat reinfections exhibited a distinction between the pre-omicron (Figure 1B) and omicron era (Figure 1C). Reinfections were rare before omicron’s emergence but became widespread during the omicron era, including among individuals previously infected with omicron. These findings are even more remarkable that the documented reinfections represent only a fraction of the actual occurrences (Section S4).

These findings may indicate accelerated viral evolution in the omicron era aimed at evading population immunity, but with minimal impact on COVID-19 severity,^2^ or potentially suggest immune imprinting effects^3–5^ that require further investigation.

## Oversight

The institutional review boards at Hamad Medical Corporation and Weill Cornell Medicine– Qatar approved this retrospective study with a waiver of informed consent. The study was reported according to the Strengthening the Reporting of Observational Studies in Epidemiology (STROBE) guidelines (Table S3). The authors vouch for the accuracy and completeness of the data and for the fidelity of the study to the protocol.

## Author contributions

HC co-designed the study, performed the statistical analyses, and co-wrote the first draft of the article. LJA conceived and co-designed the study, led the statistical analyses, and co-wrote the first draft of the article. PVC conducted viral genome sequencing and designed mass PCR testing to allow routine capture of SGTF variants. PT and MRH conducted the multiplex, real-time reverse-transcription PCR variant screening and viral genome sequencing. HY and AAA-T conducted viral genome sequencing. All authors contributed to data collection and acquisition, database development, discussion and interpretation of the results, and to the writing of the manuscript. All authors have read and approved the final manuscript.

## Data Availability

The dataset of this study is a property of the Qatar Ministry of Public Health that was provided to the researchers through a restricted-access agreement that prevents sharing the dataset with a third party or publicly. The data are available under restricted access for preservation of confidentiality of patient data. Access can be obtained through a direct application for data access to Her Excellency the Minister of Public Health (https:// www.moph.gov.qa/english/OurServices/eservices/Pages/Governmental-HealthCommunication-Center.aspx). The raw data are protected and are not available due to data privacy laws. Aggregate data are available within the paper and its supplementary information.

## Acknowledgements and support

We acknowledge the many dedicated individuals at Hamad Medical Corporation, the Ministry of Public Health, the Primary Health Care Corporation, the Qatar Biobank, Sidra Medicine, and Weill Cornell Medicine – Qatar for their diligent efforts and contributions to make this study possible.

The authors are grateful for support from the Biomedical Research Program and the Biostatistics, Epidemiology, and Biomathematics Research Core, both at Weill Cornell Medicine-Qatar, as well as for support provided by the Ministry of Public Health, Hamad Medical Corporation, and Sidra Medicine. The authors are also grateful for the Qatar Genome Programme and Qatar University Biomedical Research Center for institutional support for the reagents needed for the viral genome sequencing. Statements made herein are solely the responsibility of the authors.

The funders of the study had no role in study design, data collection, data analysis, data interpretation, or writing of the article.

## Supplementary Material

### Section S1. Further details on study methods

#### Study population and data sources

This study was conducted on the population of Qatar including data between February 28, 2020, marking the first documented severe acute respiratory syndrome coronavirus 2 (SARS-CoV-2) infection in Qatar, and June 11, 2023, end of the study. It analyzed the national, federated databases for coronavirus disease 2019 (COVID-19) laboratory testing, vaccination, hospitalization, and death, retrieved from the integrated, nationwide, digital-health information platform. Databases include all SARS-CoV-2-related data with no missing information since the onset of the pandemic, including all polymerase chain reaction (PCR) tests and medically supervised rapid antigen tests (Section S2).

Qatar’s national and universal public healthcare system uses the Cerner-system advanced digital health platform to track all electronic health record encounters of each individual in the country, including all citizens and residents registered in the national and universal public healthcare system. Registration in the public healthcare system is mandatory for citizens and residents.

The databases analyzed in this study are data-extract downloads from the Cerner-system that have been implemented on a regular (twice weekly) schedule since the onset of pandemic by the Business Intelligence Unit at Hamad Medical Corporation. Hamad Medical Corporation is the national public healthcare provider in Qatar. At every download all tests, COVID-19 vaccinations, hospitalizations related to COVID-19, and all death records regardless of cause are provided to the authors through .csv files. These databases have been analyzed throughout the pandemic not only for study-related purposes, but also to provide policymakers with summary data and analytics to inform the national response.

Every health encounter in the Cerner-system is linked to a unique individual through the HMC Number that links all records for this individual at the national level. Databases were merged and analyzed using the HMC Number to link all records whether for testing, vaccinations, hospitalizations, and deaths. All deaths in Qatar are tracked by the public healthcare system.^1, 2^ All COVID-19-related healthcare was provided only in the public healthcare system. No private entity was permitted to provide COVID-19-related hospitalization. COVID-19 vaccination was also provided only through the public healthcare system. These health records were tracked throughout the COVID-19 pandemic using the Cerner system. This system has been implemented in 2013, before the onset of the pandemic. Therefore, we had all health records related to this study for the full national cohort of citizens and residents throughout the pandemic. This allowed us to follow each person over time.

Demographic details for every HMC Number (individual) such as sex, age, and nationality are collected upon issuing of the universal health card, based on the Qatar Identity Card, which is a mandatory requirement by the Ministry of Interior to every citizen and resident in the country. Date of expiry of Qatar Identity Card is collected and updated at encounters with the public healthcare system. Data extraction from the Qatar Identity Card to the digital health platform is performed electronically through scanning techniques.

All SARS-CoV-2 testing in any facility in Qatar is tracked nationally in one database, the national testing database. This database covers all testing in all locations and facilities throughout the country, whether public or private. Every PCR test and a proportion of the facility-based rapid antigen tests conducted in Qatar, regardless of location or setting, are classified on the basis of symptoms and the reason for testing (clinical symptoms, contact tracing, surveys or random testing campaigns, individual requests, routine healthcare testing, pre-travel, at port of entry, or other).

Before November 1, 2022, SARS-CoV-2 testing in Qatar was done at a mass scale where close to 5% of the population were tested every week.^3, 4^ Based on the distribution of the reason for testing up to November 1, 2022, most of the tests in Qatar were conducted for routine reasons, such as being travel-related, and about 75% of documented infections were diagnosed not because of appearance of symptoms, but because of routine testing.^3, 4^

Starting from November 1, 2022, SARS-CoV-2 testing was substantially reduced, but still close to 1% of the population are tested every week.^5^ The distribution of the reason for testing between November 1, 2022 and June 11, 2023, showed that 48.2% of all tests were conducted for routine reasons. However, only 36.3% of infections were diagnosed because of routine testing. All testing results in the national testing database during the present study were factored in the analyses of this study.

The first large omicron wave that peaked in January of 2022 was massive and strained the testing capacity in the country.^3, 6, 7^ Accordingly, rapid antigen testing was introduced to relieve the pressure on PCR testing. Implementation of this change in testing occurred quickly precluding incorporation of reason for testing in large proportion of the rapid antigen tests for several months. While the reason for testing is available for all PCR tests, it is not available for all rapid antigen tests. Availability of reason for testing for the rapid antigen tests also varied with time.

Rapid antigen test kits are available for purchase in pharmacies in Qatar, but outcome of home-based testing is not reported nor documented in the national databases. Since SARS-CoV-2-test outcomes are linked to specific public health measures, restrictions, and privileges, testing policy and guidelines stress facility-based testing as the core testing mechanism in the population.

While facility-based testing is provided free of charge or at low subsidized costs, depending on the reason for testing, home-based rapid antigen testing is de-emphasized and not supported as part of national policy.

Qatar has unusually young, diverse demographics, in that only 9% of its residents are ≥50 years of age, and 89% are expatriates from over 150 countries.^8, 9^ Qatar launched its COVID-19 vaccination program in December of 2020 using BNT162b2 and mRNA-1273 vaccines,^10^ and initiated vaccination with the 50-μg bivalent mRNA-1273.214 vaccine^11^ in October of 2022.^5^ These vaccines are accessible at multiple facilities throughout the country and are provided free charge regardless of citizenship or residency status. Further descriptions of the study population and these national databases were reported previously.^1–4, 7, 9, 12, 13^

#### Statistical analysis

Primary infections and reinfections were described using frequency distributions and measures of central tendency. Primary infection was defined as the first recorded instance of a positive SARS-CoV-2 test for an individual. Reinfection was defined as a documented positive SARS-CoV-2 test occurring at least 90 days after a previous documented infection. This definition was implemented to prevent misclassifying prolonged SARS-CoV-2 positivity^14^ as reinfection.^6, 15, 16^

The proportion of primary infections on a given day was determined by dividing the number of infections diagnosed on that day that were classified as primary infections by the total number of infections diagnosed on that day. The proportion of first reinfections on a given day was determined by dividing the number of infections diagnosed on that day that were classified as first reinfections by the total number of infections diagnosed on that day. The proportion of second (or higher order) reinfections was defined in a similar manner.

Statistical analyses were performed using Stata/SE version 17.0 (Stata Corporation, College Station, TX, USA).

### Section S2. Laboratory methods and variant ascertainment

#### Real-time reverse-transcription polymerase chain reaction testing

Nasopharyngeal and/or oropharyngeal swabs were collected for polymerase chain reaction (PCR) testing and placed in Universal Transport Medium (UTM). Aliquots of UTM were: 1) extracted on KingFisher Flex (Thermo Fisher Scientific, USA), MGISP-960 (MGI, China), or ExiPrep 96 Lite (Bioneer, South Korea) followed by testing with real-time reverse-transcription PCR (RT-qPCR) using TaqPath COVID-19 Combo Kits (Thermo Fisher Scientific, USA) on an ABI 7500 FAST (Thermo Fisher Scientific, USA); 2) tested directly on the Cepheid GeneXpert system using the Xpert Xpress SARS-CoV-2 (Cepheid, USA); or 3) loaded directly into a Roche cobas 6800 system and assayed with the cobas SARS-CoV-2 Test (Roche, Switzerland). The first assay targets the viral S, N, and ORF1ab gene regions. The second targets the viral N and E-gene regions, and the third targets the ORF1ab and E-gene regions.

All PCR testing was conducted at the Hamad Medical Corporation Central Laboratory or Sidra Medicine Laboratory, following standardized protocols.

#### Rapid antigen testing

Severe acute respiratory syndrome coronavirus 2 (SARS-CoV-2) antigen tests were performed on nasopharyngeal swabs using one of the following lateral flow antigen tests: Panbio COVID-19 Ag Rapid Test Device (Abbott, USA); SARS-CoV-2 Rapid Antigen Test (Roche, Switzerland); Standard Q COVID-19 Antigen Test (SD Biosensor, Korea); or CareStart COVID-19 Antigen Test (Access Bio, USA). All antigen tests were performed point-of-care according to each manufacturer’s instructions at public or private hospitals and clinics throughout Qatar with prior authorization and training by the Ministry of Public Health (MOPH). Antigen test results were electronically reported to the MOPH in real time using the Antigen Test Management System which is integrated with the national Coronavirus Disease 2019 (COVID-19) database.

#### Classification of infections by variant type

Surveillance for SARS-CoV-2 variants in Qatar is based on viral genome sequencing and multiplex RT-qPCR variant screening^17^ of random positive clinical samples,^4, 18–22^ complemented by deep sequencing of wastewater samples.^20, 23, 24^ Further details on the viral genome sequencing and multiplex RT-qPCR variant screening throughout the SARS-CoV-2 waves in Qatar can be found in previous publications.^3, 4, 6, 13, 18–22, 25–29^

### Section S3. Phases of the COVID-19 pandemic in Qatar

Throughout the study period, SARS-CoV-2 incidence was initially driven by the ancestral virus. However, over time, a series of variants came to dominate the incidence including alpha, beta, delta, omicron BA.1 and BA.2, omicron BA.4 and BA.5, omicron BA.2.75*, and finally omicron XBB*.

The pandemic was categorized into distinct phases based on the level of SARS-CoV-2 incidence and the predominant variant. These phases included the ancestral virus wave (February 28, 2020 - July 31, 2020),^9^ a prolonged low incidence phase with the ancestral virus (August 1, 2020 - January 17, 2021),^4, 30^ the alpha wave (January 18, 2021 - March 7, 2021),^31^ the beta wave (March 8, 2021 - May 31, 2021),^12^ a prolonged low incidence delta phase (June 1, 2021 - December 18, 2021),^25, 32^ the first (BA.1 & BA.2) omicron wave (December 19, 2021 - February 28, 2022),^33^ the omicron BA.4 & BA.5 wave (March 1, 2022 - September 9, 2022),^28^ and the omicron BA.2.75 & XBB waves (September 10, 2022 - June 11, 2023).^5^

### Section S4. Limitations and caveats

This study has limitations that may lead to an underestimation of the numbers and proportions of first (or higher order) reinfections. Firstly, the analysis relied on documented infections, potentially overlooking unrecorded infections in individuals who were asymptomatic or not tested. Secondly, the operational definition used in this study for reinfection excluded reinfections occurring within 90 days of a previous infection, which may have resulted in the omission of some reinfection cases.

Thirdly, testing frequency varies among different sectors of the population, leading to variations in the likelihood of documenting infections. Some sectors may undergo more frequent testing, increasing the chances of detecting and documenting infections and reinfections, while others may have lower testing frequencies, reducing the likelihood of capturing infections and reinfections.

Fourthly, this study focused exclusively on documented infections in Qatar, which has a distinctive demographic composition. With 89% of the population comprising expatriates from over 150 countries,^8, 9^ there is possibly a higher likelihood of travel within this population compared to other national populations. As a result, it is possible that some infections occurred while individuals were abroad and thus were not recorded in the national COVID-19 databases, leading to their exclusion from our analysis.

Given our definition of reinfection as a documented positive SARS-CoV-2 test occurring at least 90 days after a previous documented infection, it is possible that prolonged infections^14^ lasting longer than 90 days could have been misclassified as reinfections.^6, 15, 16^ However, occurrence of such prolonged infections among the healthy and young population in Qatar is expected to be minimal.

The observed proportions of reinfections appeared to plateau from November 2022 onwards. However, this pattern may be influenced by the significant reduction in testing that occurred during this month, leading to a decreased ability to identify and capture reinfections. The actual proportions of reinfections may have continued to increase, but the under-detection of reinfections could have hindered our ability to observe this trend.

Differences in the distributions of primary infections, first reinfections, and higher order reinfections were observed across eras (pre-omicron versus omicron), age groups, sexes, nationalities, and number of coexisting conditions (Tables S1 and S2). These differences may have arisen from variations in the population sector impacted by each wave and differences in testing rates. Other factors that remain to be clarified may have also contributed to these differences.

With the relatively young population of Qatar, our findings may not be generalizable to other countries where elderly citizens constitute a larger proportion of the total population.

---

**Table S1.**
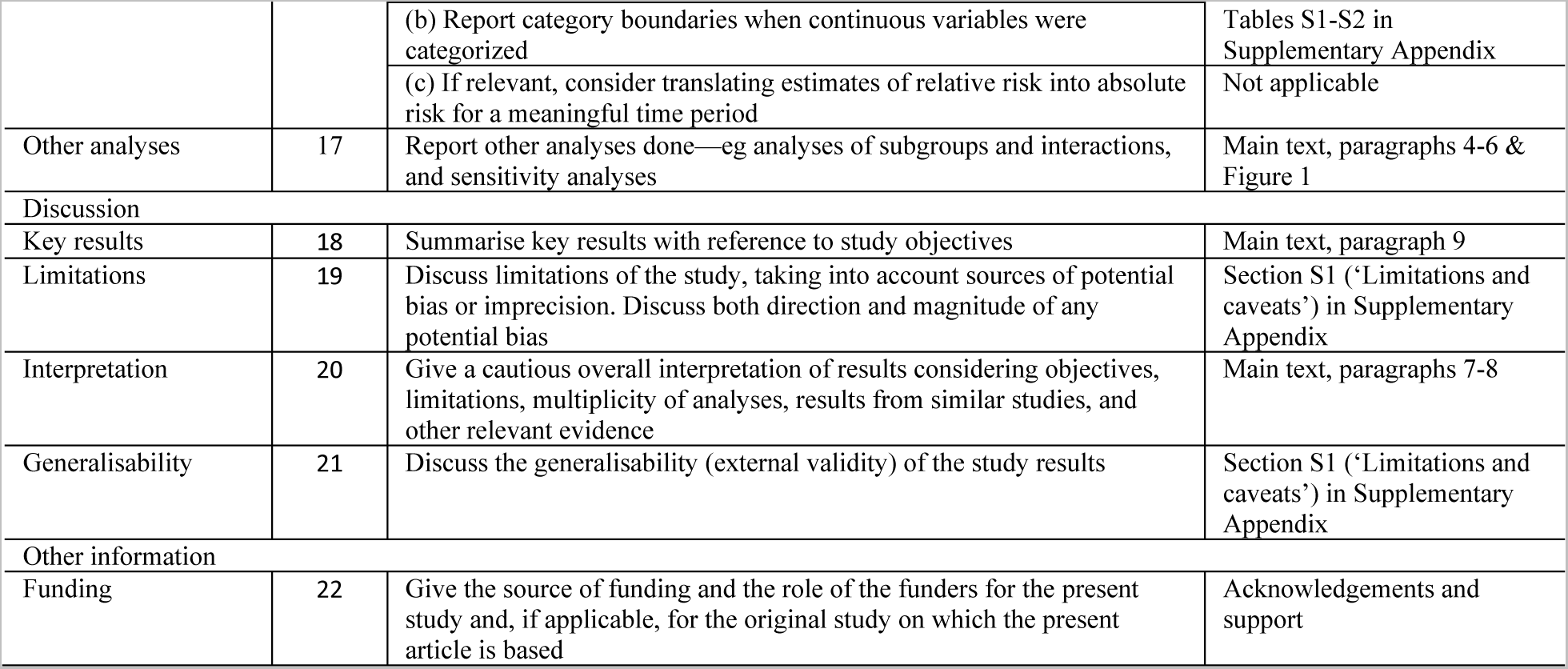
Characteristics of SARS-CoV-2 primary infections, first reinfections, and second reinfections in the pre-omicron era.

**Table S2.**
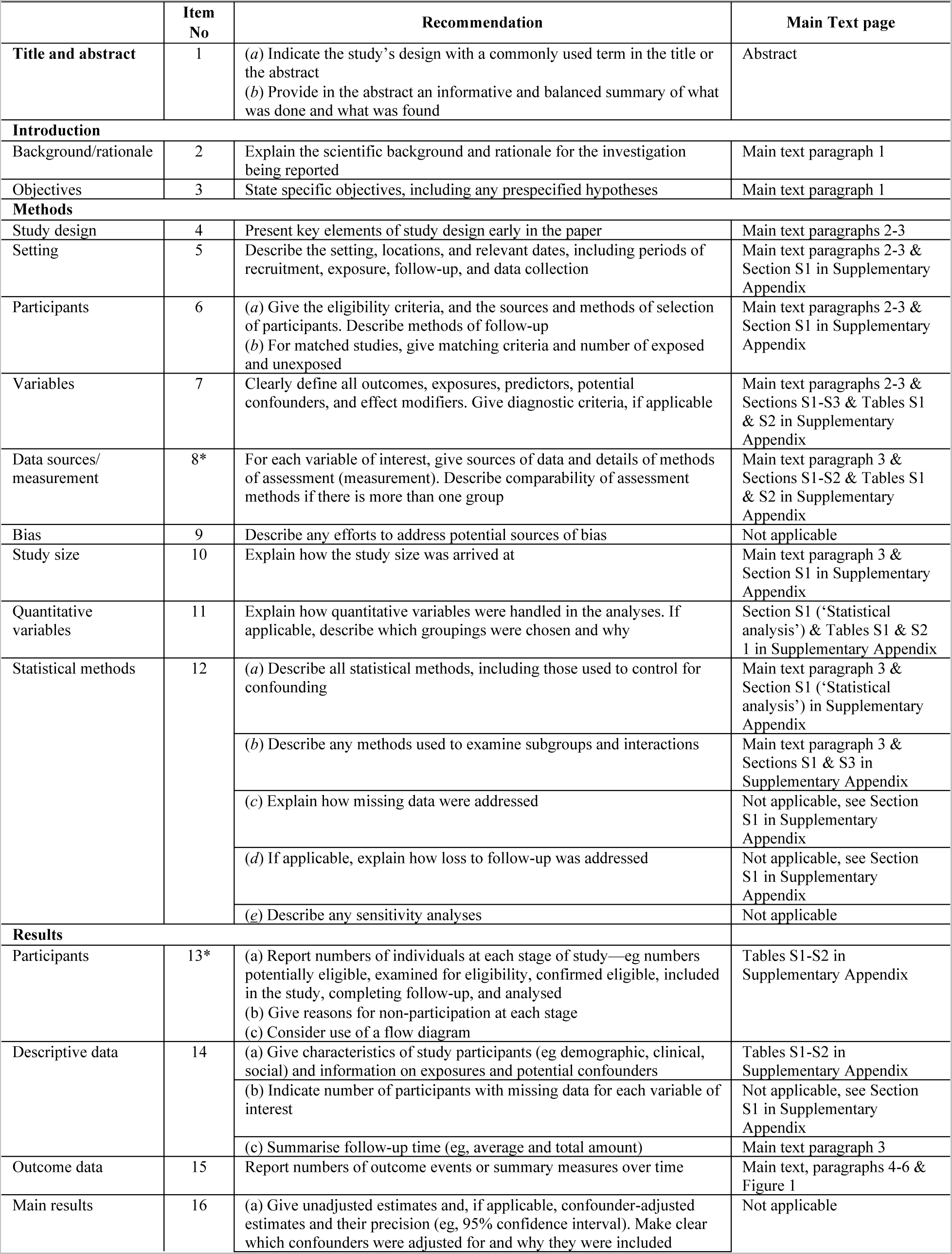
Characteristics of SARS-CoV-2 primary infections, first reinfections, second reinfections, and third reinfections in the omicron era.

**Table S3.**
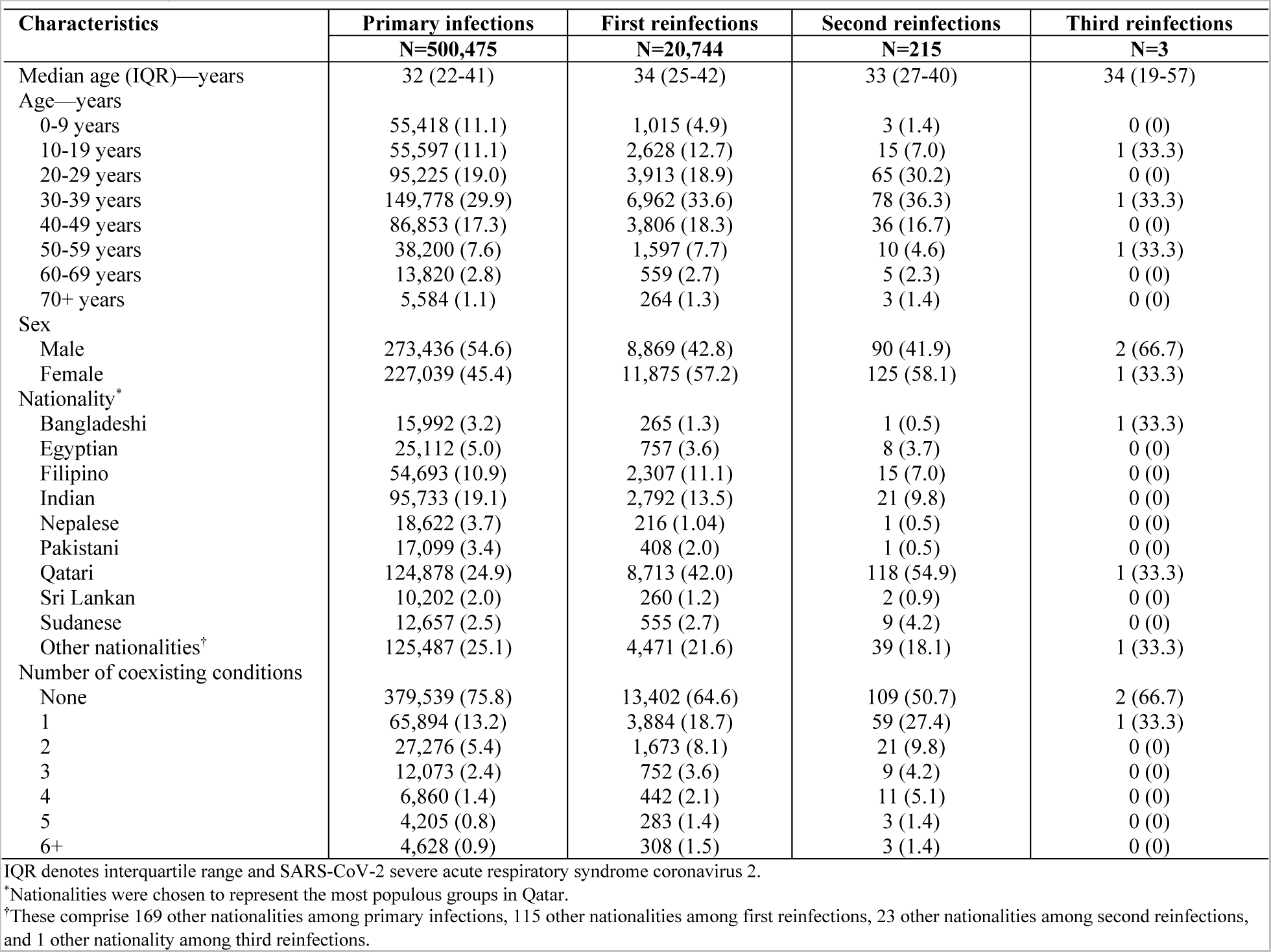
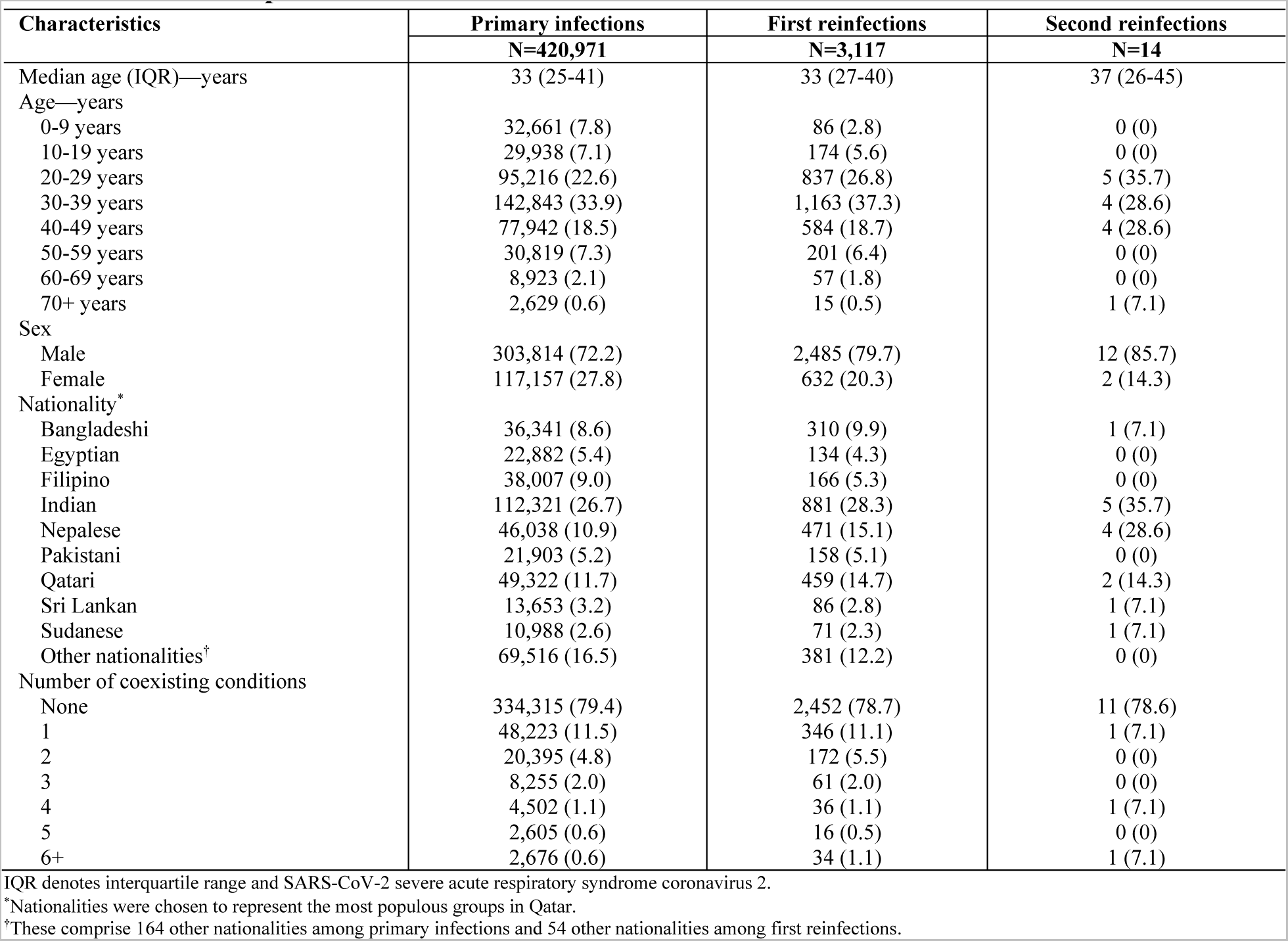
STROBE checklist for cohort studies.

